# Diagnostic Performance of Systemic-Immune Inflammation Index for Overall and Complicated Acute Appendicitis: A Systematic Review and a Diagnostic Test Accuracy Meta-Analysis

**DOI:** 10.1101/2024.09.24.24314119

**Authors:** Javier Arredondo Montero, Carlos Delgado-Miguel, Blanca Paola Pérez Riveros, Rafael Fernández-Atuan, María Rico-Jiménez

**Author notes:** <u>Corresponding author:</u> Javier Arredondo Montero, MD, PhD, Adress: Department of Pediatric Surgery, Complejo Asistencial Universitario de León. c/Altos de Nava s/n, 24008 León, Castilla y León, Spain, /. **CRediT authorship contribution statement**: **JAM:** Conceptualization and study design; literature search and selection; data curation and extraction; formal analysis; investigation; methodology; project administration; resources; validation; visualization; writing – original draft; writing – review and editing. **MRJ:** literature search and selection; data curation and extraction; writing – review and editing. **BPR**: literature search and selection; data curation and extraction. **CDM, RFA:** writing – review and editing. **CONFLICTS OF INTEREST:** The authors declare that they have no conflict of interest. **FINANCIAL STATEMENT/FUNDING:** This review did not receive any specific grant from funding agencies in the public, commercial, or not-for-profit sectors, and none of the authors has external funding to declare. **ETHICAL APPROVAL:** This study did not involve the participation of human or animal subjects, and therefore, IRB approval was not sought. **STATEMENT OF AVAILABILITY OF THE DATA USED DURING THE SYSTEMATIC REVIEW:** The data used to carry out this systematic review are available upon request from the authors.

## Abstract

**Background:** This study aimed to analyze the systemic-immune inflammation index (SII)’s diagnostic performance in diagnosing acute appendicitis (AA) and discriminating between complicated acute appendicitis (CAA) and non-complicated acute appendicitis (NCAA).

**Methods:** This review was registered in PROSPERO (CRD42024587430). We included prospective or retrospective original clinical studies evaluating the diagnostic performance of SII in AA. A search was conducted in PubMed, Web of Science, Scopus, and OVID. Search terms and keywords were: (appendicitis OR appendectomy) AND (systemic-immune inflammation index OR SII). Two independent reviewers selected the articles and extracted relevant data. Methodological quality was assessed using the QUADAS2 index. A synthesis of the results, standardization of the metrics, four random-effect meta-analyses, and two Diagnostic Test Accuracy (DTA) meta-analyses were performed.

**Results:** Thirteen studies with data from 9083 participants, including 5255 patients with a confirmed diagnosis of AA and 3828 controls (CG), were included in this review. The random-effect meta-analysis of SII (AA vs. CG) included ten articles (3733 AA and 3510 controls) and resulted in a significant mean difference [95% CI] of 1072.46 [750.55,1394.37] (p<0.001). The random-effect meta-analysis of SII (CAA vs. NCAA) included nine articles (1116 CAA and 2984 NCAA) and resulted in a significant mean difference [95% CI] of 1294.2 [731.54,1856.86] (p<0.001). Subgroup meta-analysis for studies conducted in pediatric-only populations maintained statistical significance. The DTA meta-analysis (AA vs. CG) yielded a pooled sensitivity and specificity [95% CI] of 81.8 [75.2,86.9] % and 79.9 [68.2,88.1] %. The DTA meta-analysis (NCAA vs. CAA) resulted in a pooled sensitivity and specificity [95% CI] of 72.5 [49.6,87.6] % and 82.5 [65.1,92.2] %.

**Conclusions:** SII emerges as a robust tool for diagnosing AA and differentiating between NCAA and CAA. The retrospective nature of most of the included studies and their limited geographical distribution warrant further prospective multicenter studies to validate these findings.

**Funding:** None

**Registration:** PROSPERO (CRD42024587430).

## Introduction

Acute appendicitis (AA), the most common emergency surgical pathology worldwide [1], remains a pathology with a significant misdiagnosis rate [2]. In recent years, multiple lines of research have been developed to optimize the diagnostic performance of this entity. On the one hand, numerous biomarkers have been assessed in different biological samples, such as peripheral blood [3] and urine [4]. Nevertheless, most of the molecules studied have achieved only moderate diagnostic yields, which excludes their use as a single diagnostic test for this pathology [3–4]. In addition, these molecules often require lengthy processing times and involve high economic and human costs, which limits their potential implementability in clinical practice [5]. Concerning imaging tests in AA, recent meta-analyses demonstrate very high sensitivity and specificity for computed tomography (CT) [6]. Still, it must be considered that it is a radiological study that implies high radiation exposure. Its use is restricted to particular situations in specific population subgroups, such as pediatrics [7]. Likewise, the economic cost and logistical complexity of CT are high, so it cannot be considered a routine test in all patients with suspected acute appendicitis. On the other hand, ultrasound (US) is a widely available, inexpensive, and rapid test that is not associated with ionizing radiation. Still, its overall diagnostic performance is considerably worse, with 82% and 86% pooled sensitivities and specificities, respectively [6]. Although recent studies address exciting and novel aspects concerning the role of imaging tests in the diagnosis of AA, such as the role of point-of-care ultrasound [8] or the assessment of specific ultrasound parameters such as appendiceal vascularization [9], none of these approaches has demonstrated superior diagnostic performance to pre-existing ones. MRI has also shown great potential for diagnosing AA and is not associated with ionizing radiation [10]. Still, the need for specialized personnel and equipment and the economic and processing costs hinder its implementability as a routine tool for diagnosing AA. Finally, other recent lines of research, such as the use of artificial intelligence (AI) diagnostic models, also show potential but are still in preliminary stages and require development and external validation [11].

Hemogram-derived ratios are dimensionless parameters derived from various mathematical formulae involving the main elements of the complete blood count (CBC). Prominent examples are the neutrophil-to-lymphocyte ratio (NLR), the platelet-to-lymphocyte ratio (PLR), and the lymphocyte-to-monocyte ratio (LMR), which have demonstrated diagnostic and prognostic utility in many pathologies, including AA [12–15].

Recently, more complex indices have been described, such as the systemic-immune inflammatory index (SII), which is the result of multiplying the total neutrophil count (TNC) by the total platelet count (TPC) and dividing the result by the total lymphocyte count (TLC) (TNC x TPC / TLC). SII, which has also proven helpful in multiple pathologies [16–18], is of particular interest in acute inflammatory or infectious pathologies due to the biological plausibility of its approach: marked neutrophilia, reactive thrombocytosis (platelets acting as an acute phase reactant) and lymphopenia secondary to marked neutrophilia are common in these patients.

The present work aims to evaluate the SII capacity for diagnosing AA in adult and pediatric populations and its discriminative capacity between complicated appendicitis (CAA) and non-complicated appendicitis (NCAA).

## Methods

### Literature search and selection

We followed the Preferred Reporting Items for Systematic Reviews and Meta-Analyses in Diagnostic Test Accuracy Studies (PRISMA-DTA) guidance [19]. We specifically designed and implemented a review protocol registered in the International Prospective Register of Systematic Reviews (PROSPERO ID CRD42024587430). Eligible studies were identified by searching the primary existing medical bibliography databases (PubMed, Web of Science, Scopus, and OVID). Search terms and keywords were: (appendicitis OR appendectomy) AND (systemic-immune inflammation index OR SII). The search was last executed on 15.09.2024.

Supplementary File 1 shows inclusion and exclusion criteria. JAM and BPR selected articles using the COVIDENCE ® tool. The search results were imported into the platform, and both authors screened the articles separately. Disagreements were resolved by consensus.

### Quality assessment

The QUADAS2 tool was used to evaluate each selected articlés methodological quality and risk of bias [20]. Each article evaluated patient selection, index test, reference standard, and flow and timing. Applicability concerns regarding patient selection, index tests, and reference standards were also assessed.

### Data extraction and synthesis

The target condition was defined as histopathologically confirmed AA or formal clinical suspicion of AA. The index test was defined as SII. The reference standard was defined as the histopathological study of the cecal appendix. Three independent reviewers (JAM, MRJ, BPR) extracted the relevant data from the selected articles following a standardized procedure.

Extracted data included author, country where the study was conducted, year of publication, study design, study population (sample size, age range, and sex distribution), AA group and control group (CG) definitions, reference standard used in AA group, mean and standard deviation (or median and interquartile range) for SII, statistical p-value for the between-group comparison, SII area under the curve (AUC), SII cut-off value (if established), and its associated sensitivity and specificity. There were no disagreements between the reviewers after collating the extracted data. The metrics used in each study were reviewed, and a standardization of units was performed in one case. Medians (Interquartile ranges) were converted to means (standard deviations) following a standardized procedure [21]. True positives (TP), false positives (FP), true negatives (TN), and false negatives (FN) values were calculated for all articles that provided sufficient data using standardized statistical formulae [22]. The reported predicted values were used to check that the calculations were performed correctly.

### Meta-analysis

Four random-effects meta-analyses (MA) for SII were performed using the restricted maximum likelihood method (REML): 1) MA for SII values (AA vs. CG), including all studies with available data except Saridas et al. and Guo et al. 2) MA for SII values (CAA vs. NCAA), including all studies with available data except Ertekin et al., Duyan et al., Şener et al. and Guo et al. 3) MA for SII values (AA vs. CG), including only studies with an exclusive pediatric population. 4) MA for SII values (CAA vs. NCAA), including only studies with an exclusive pediatric population. Additional sensitivity analyses were performed to explore potential sources of heterogeneity. The results were expressed as a mean difference (95% CI) and were plotted in a forest plot. Between-study heterogeneity was assessed using the I^2^ statistics. Additionally, two leave-one-out random-effects MA were performed (one for MA 1 and one for MA 2).

### Diagnostic Test Accuracy Meta-analysis

Two diagnostic test accuracy (DTA) meta-analytical models were performed: 1) SII diagnostic performance (AA vs. CG) and 2) SII discriminative ability (CAA vs. NCAA). Pooled sensitivity and specificity were reported for each of the meta-analytical models. The results were depicted as forest plots of sensitivities and specificities and hierarchical summary receiver operating characteristic (hSROC) curves.

### Publication Bias and Small-Study Effects Assessment

Egger’s test and funnel plots (not shown) were used to assess the risk of publication bias. The trim-and-fill method for publication bias was employed in cases where potential publication bias was identified [23].

## Results

The search returned 63 articles (Scopus n=4; Pubmed n=16; Web of Science n=29; OVID n=14). Twenty-eight duplicates were removed. Among the remaining 35 articles, we excluded 22 (inclusion and exclusion criteria, n=22; reports not retrieved, n=0). This review finally included 13 studies with data from 9083 participants, including 5255 patients with a confirmed diagnosis of AA and 3828 controls (CG) [24–36]. The flowchart of the search and selection process is shown in Figure 1.

**Figure 1.**
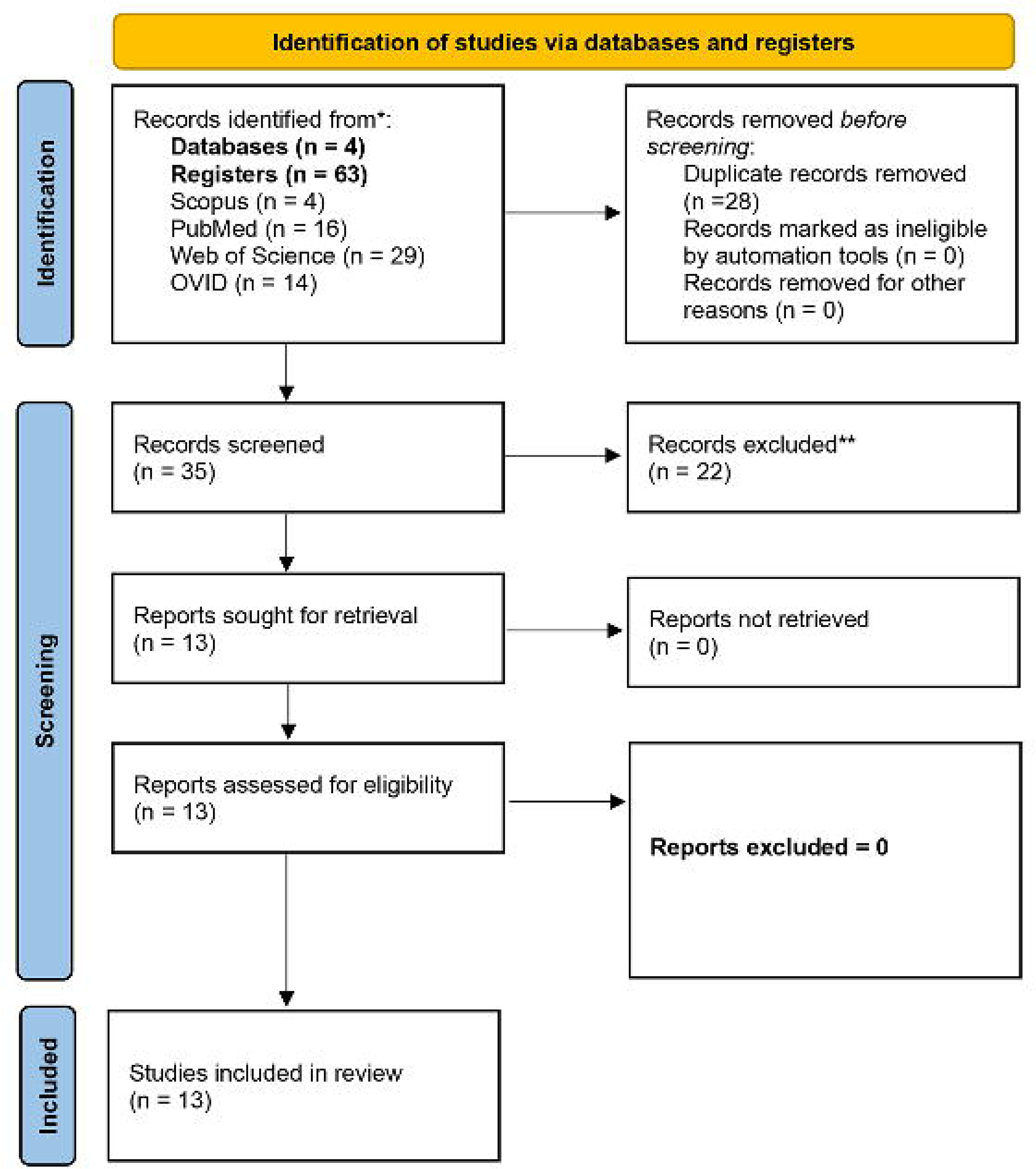
Flowchart of the search and selection process.

The risk of bias concerning the selection of patients was considered low in one of the studies [27], unclear in ten of them [24–26,28–31,33,34,36], and high in two of them [32,35]. The risk of bias concerning the index test was considered low in twelve studies [24–31,33–36] and high in one [32]. The risk of bias concerning the reference standard was considered low in all the studies [24–36]. The risk of bias concerning flow and timing was considered low in six studies [27,30,31,33,34,35] and unclear in seven studies [24,25,26,28,29,32,36]. Regarding patient selection applicability concerns, the risk was considered low in ten of the studies [24,25,27,28,29,30,31,33,34,36], unclear in one of the studies [26], and high in two of them [32,35]. Regarding index test applicability concern, the risk was considered low in twelve studies [24–31,33–36] and high in one study [32]. Concerning reference standard applicability concerns, the risk was considered low in twelve studies [24–34,36] and high in one study [35]. The results of the QUADAS2 analysis are shown in Figure 2.

**Figure 2.**
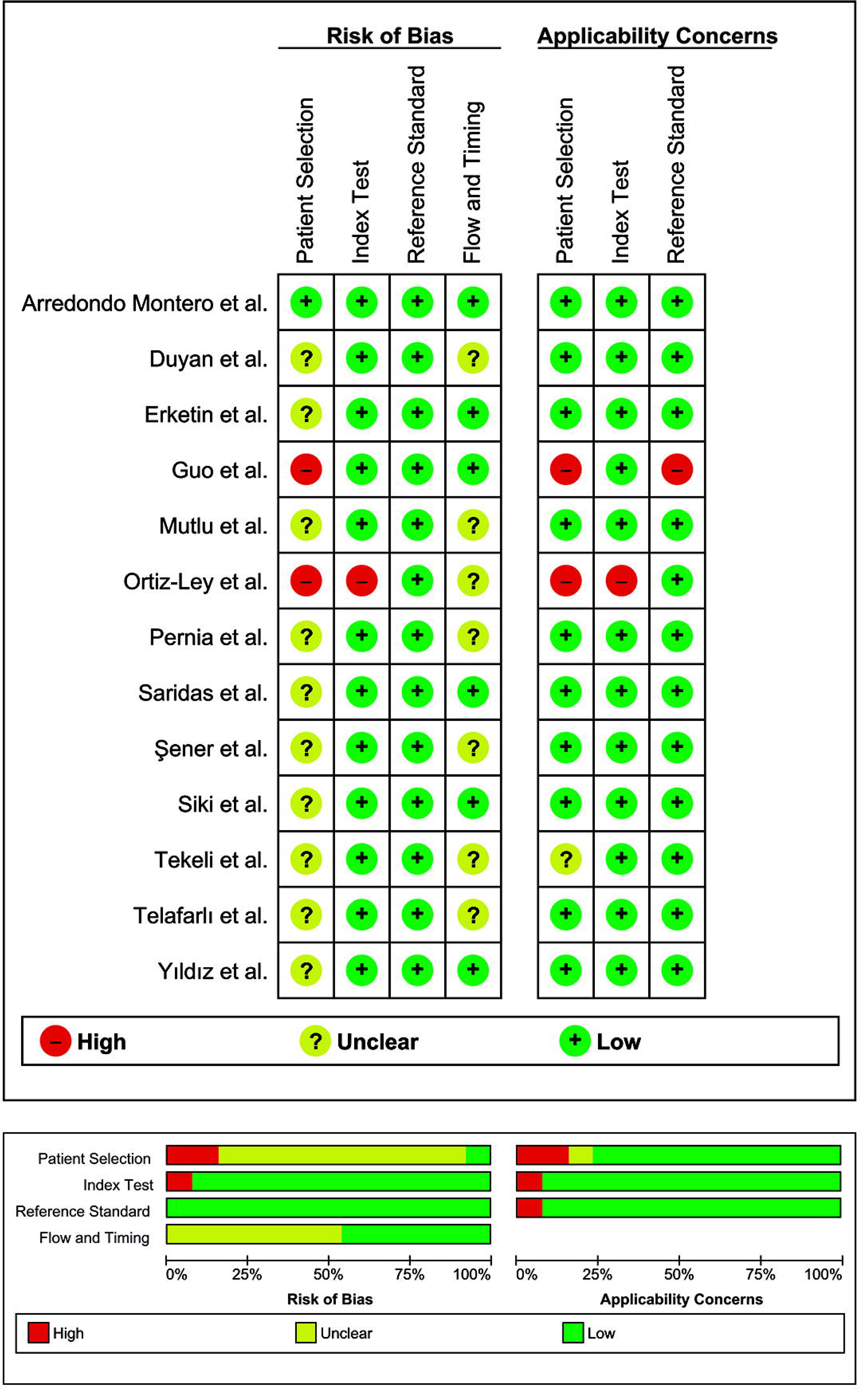
Graphical representation of the quality assessment of the diagnostic accuracy studies included in the review (QUADAS2).

## Systemic-Immune Inflammation Index in Acute Appendicitis

### Sociodemographic Characteristics

Table 1 summarizes the data extracted from the thirteen studies that evaluated SII. All studies were carried out between 2022 and 2024. Two were from Spain [27,36], one was from China [35], one was from Mexico [32], and nine were from Turkey [24–26,28–31,33,34]. One study was prospective [27], and twelve were retrospective [24–26,28–36]. Six studies involved pediatric populations [26,27,31,32,35,36].

“The definition of “case” was consistent in all the selected studies, given as the histopathological confirmation of AA in the surgical specimen [24–36]. Eleven studies stratified the AA group into NCAA and CAA [25–34,36].

This was not the case for the definition of “control,” which constituted either patients seen at the Emergency Department in which the diagnosis of AA was finally excluded (also known as non-surgical abdominal pain or NSAP) [24,25,27,29], patients seen at the Emergency Department with non-inflammatory abdominal pain (NIAP) (renal colic, inguinal hernia, umbilical hernia, etc.) [24,28], patients admitted to the outpatient clinic for umbilical hernia, inguinal hernia, and circumcision [26], negative appendectomies (NA) [30,31], patients seen in outpatient presurgical consultation without any inflammatory or infectious process [32], patients with chronic appendicitis [35] and patients with suspected AA who went to the Emergency Department did not underwent surgery during that episode (considering the episode of one week’s duration from the first visit to the Emergency Department) [36].

Concerning exclusion criteria, they were as follows: pregnant women [24,29,30,33], patients with heart failure [24,33,35], patients with hematological disease [24,27,28,29,30,32,33,34,35] or liver disease [24,28,32,33,34,35], patients with kidney failure [30,33,35], patients with mental or cognitive impairments [35], patients with peripheral vascular disease [24], patients using anticoagulants, antibiotics, chemotherapy and/or steroids or immunomodulators [24,27,30,32,33,34], patients with acute infections [24,28,32,34,35] or chronic infections [24,32,34,35], patients with fever of unknown origin [28], patients with appendiceal neoplasms as a result of the histological analysis of the surgical specimen [24,28,34], patients with neoplasms [29,32,33,35] or metastatic neoplasms [27], patients with chronic diseases [26], patients with drug use [26], patients with previous appendectomy [26,27], patients with a clear suspicion of AA or with clinical instability in which complementary tests were not performed before surgery [27], patients with previous abdominal trauma [27,33], patients with priory surgery within the last month [33], patients with inflammatory bowel disease [28], septic patients [29,30], unstable or shocked patients [29], patients diagnosed with any condition other than AA after the pathological examination [29], patients with a normal appendix in the surgery or in the pathology report [33], patients with rheumatologic or immune disorders [27,32,33,35], patients referred to an external center [30,32], patients who received medical treatments prior to admission [35] and patients with missing data [24,28,32,33,34]. Three studies did not specify their inclusion and exclusion criteria [25,31,36].

Table 1 shows the main characteristics of the studies included in this review. In some cases, discrepancies in the sample size of the studies’ subgroups were identified, and the corresponding authors were contacted for clarification. The legend in Table 1 details this.

### SII Measurement Units

Seven studies reported the values as a mean (standard deviation) [24–26,28,29,34,35] and five studies as a median (interquartile range) [27,30,31,32,33]. Pernia et al. did not provide numerical SII values. We contacted the corresponding author, and they provided us with the SII values for all groups as mean and standard deviation.

Although Ertekin et al. values were reported in the original article as median (IQR), values are more compatible with median (range). For example, the interquartile range proposed by these authors for the CAA group is 16022 (4641-773696), more than 100 times higher than other articles for the CAA group. We contacted the authors to clarify this aspect, but they did not respond. We managed them as median (IQR). Following a standardized procedure, we converted the median (range) and median (interquartile range) data to mean (standard deviation) [21].

SII is the result of a quotient and is dimensionless. In twelve articles, it was reported in a range compatible with the international system (i.e., having performed the mathematical operation with TNC, TPC, and TLC values reported in 1×10^9^/L). In the case of Ortiz-Ley et al., the units were not concordant to the other articles. Although they reported TNC, TPC, and TLC in 10^3^/µL (the same as 10^9^/L), they reported SII in x10^6^ units. The most plausible conversion was multiplying by 1×10^3^, and the values thus presented biological plausibility. The authors confirmed that this was correct.

## Systemic-Immune Inflammation Index (AA Vs. CG)

Eleven studies compared an AA group with a control group [24–32,35,36].

### Diagnostic Performance of SII (AA Vs. CG)

Nine studies provided a p-value for comparing SII values in the AA and control groups, all statistically significant [24–31,35]. One study performed a three-group comparison (CAA, NCAA, and CG) through a Kruskal-Wallis test, which was statistically significant [32].

The reported area under the curve (AUC) ranged from 0.69 [31] to 0.97 [32], and the proposed cut-offs ranged from 840.13 [25] to 4970 [32]. The reported sensitivity ranged from 68% [31] to 92% [32], and the reported specificity ranged from 61% [31] to 92% [32].

### Random-Effects Meta-Analysis for SII (AA Vs. CG)

The random-effect meta-analysis of SII (AA vs. CG) included ten articles (3733 AA and 3510 controls) and resulted in a significant mean difference [95% CI] of 1072.46 [750.55,1394.37] (p<0.001). The I^2^ value was 95.2%. A leave-one-out analysis was performed, carrying out iterations on the present model, excluding one of the studies included in the model in each iteration. The leave-one-out analysis showed that the article that conditioned the model most negatively was Ortiz-Ley et al. [32]. Its exclusion from the model resulted in a mean difference [95% CI] of 1163.35 [889.1, 1437.61]. In contrast, the article that conditioned the model most positively was Tekeli et al. Its exclusion from the model resulted in a mean difference [95% CI] of 1001.1 [856.82,1145.38]. The forest plot of this meta-analysis is shown in Figure 3.

**Figure 3.**
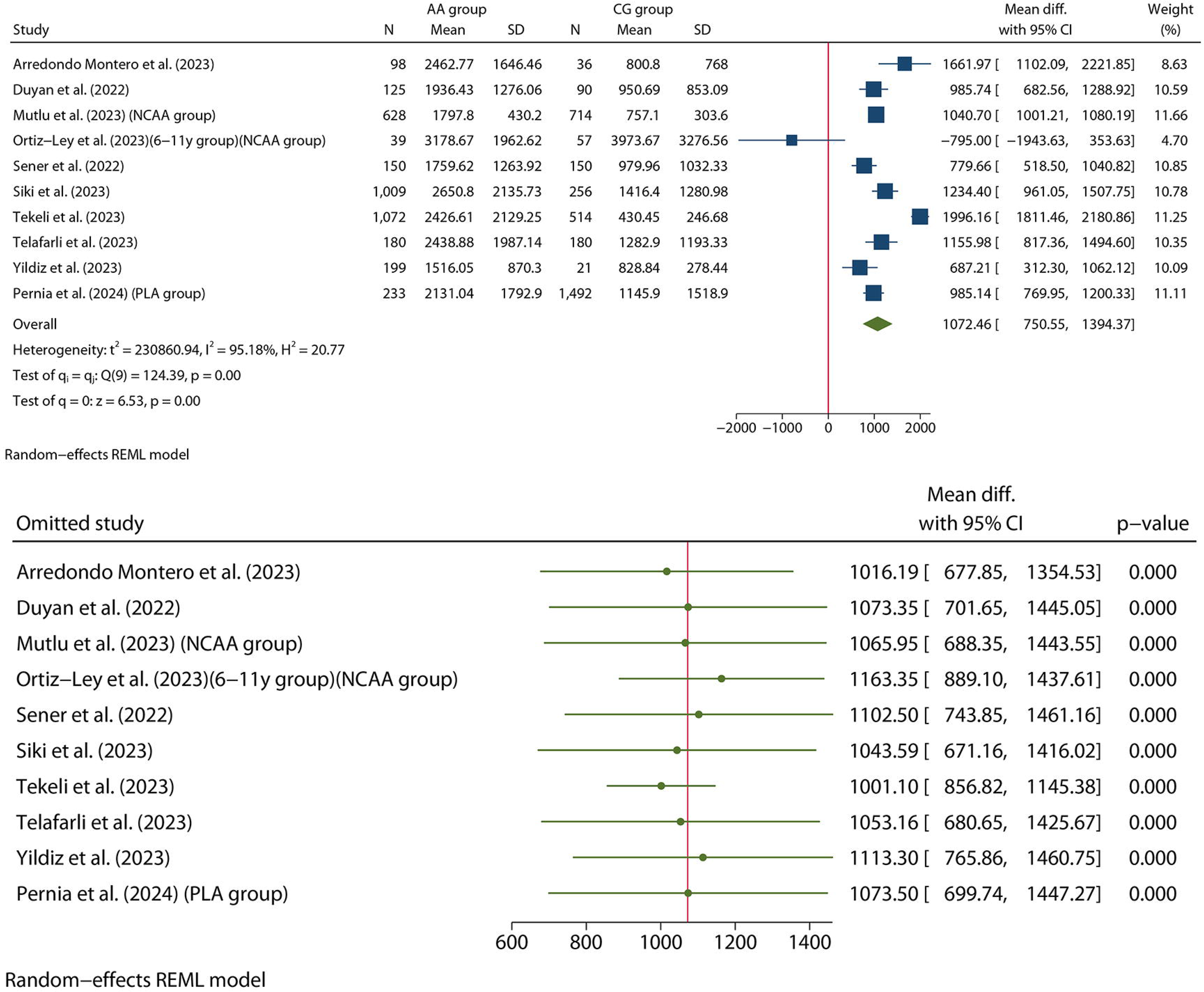
<u>Above</u>: Forest plot of the REML random-effects meta-analyses performed for SII (AA vs. CG). <u>Bottom</u>: Forest plot of the leave-one-out meta-analysis performed for SII (AA vs. CG).

An Egger test obtained a p-value of 0.04. Therefore, a trim-and-fill analysis was performed. The mean difference [95% CI] obtained was 1072.46 [750.55, 1394.37], the same as in the original model (the model did not perform any imputation).

## Systemic-Immune Inflammation Index (Complicated Appendicitis vs. Non-Complicated Appendicitis)

Ten studies compared the CAA and NCAA groups [25,26,27,28,29,30,31,32,33,34].

### Diagnostic Performance of SII (NCAA vs. CAA)

Eight studies provided a p-value for comparing SII values in the CAA and NCAA, all statistically significant [26,27,28,29,30,31,33,34]. One study performed a three-group comparison (CAA, NCAA, and CG) through a Kruskal-Wallis test, which was statistically significant [32].

The reported area under the curve (AUC) ranged from 0.595 [31] to 0.99 [33], and the proposed cut-offs ranged from 1239.77 [29] to 7560 [32]. The reported sensitivity ranged from 35.5% [27] to 99.2% [33], and the reported specificity ranged from 54% [31] to 99.5% [33].

We decided to exclude Ertekin et al. [33] from the meta-analysis because the extreme ranges they reported were inconsistent with previous reports.

The random-effect meta-analysis of SII (CAA vs. NCAA) included nine articles (1116 CAA and 2984 NCAA) and resulted in a significant mean difference [95% CI] of 1294.2 [731.54,1856.86] (p<0.001). The I^2^ value was 97.3%. The leave-one-out analysis showed that the article that conditioned the model most positively was Telafarlı et al. [29]. Their exclusion from the model resulted in a mean difference [95% CI] of 973.14 [706.92,1239.36]. The forest plot of this meta-analysis is shown in Figure 4.

**Figure 4.**
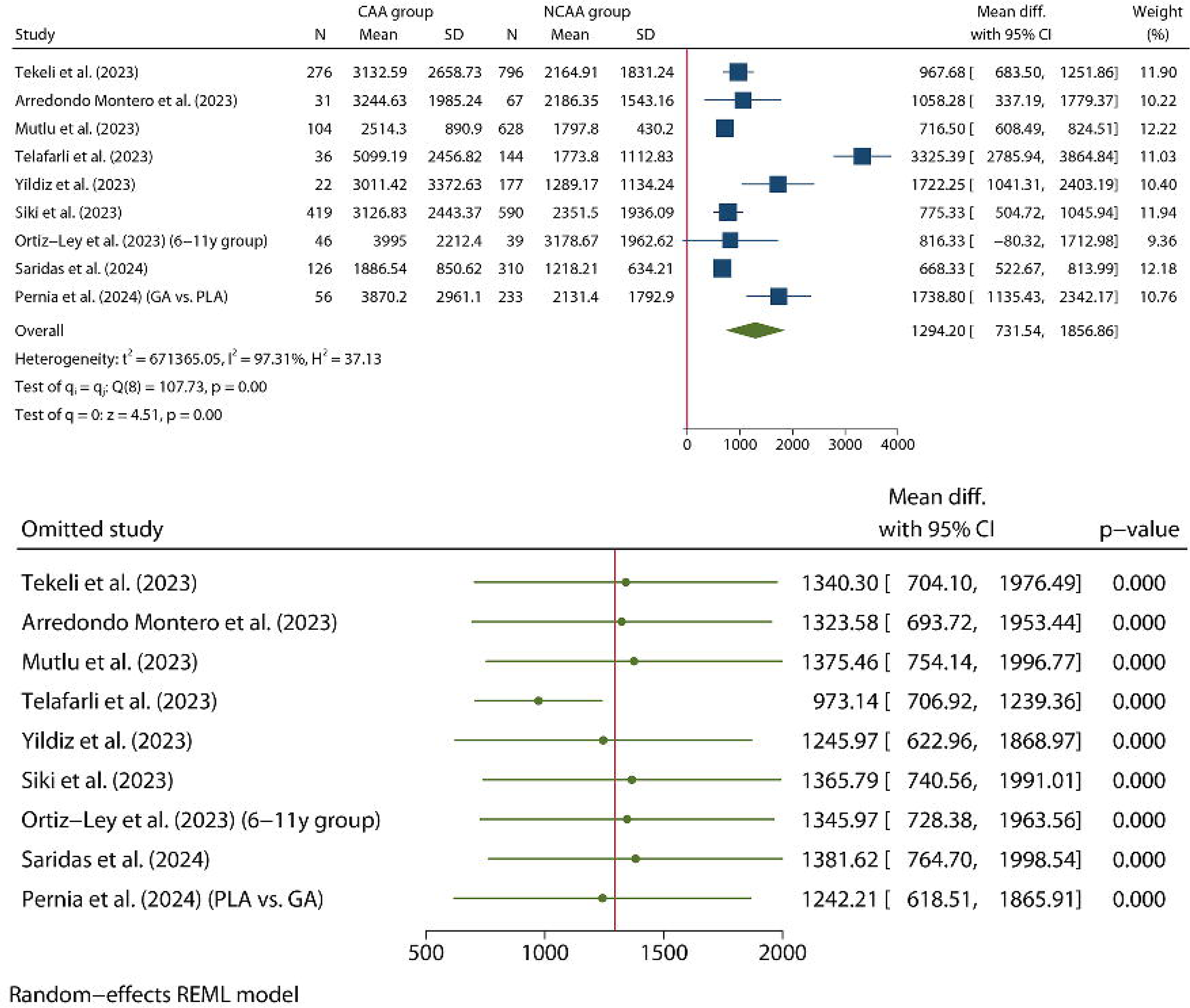
<u>Above</u>: Forest plot of the REML random-effects meta-analyses performed for SII (CAA vs. NCAA). <u>Bottom</u>: Forest plot of the leave-one-out meta-analysis performed for SII (CAA vs. NCAA).

An Egger test was performed, obtaining a p-value of 0.30. Therefore, no trim-and-fill analysis was performed.

## Systemic-Immune Inflammation Index in Pediatric Populations

Six studies were conducted on pediatric populations [26,27,31,32,35,36].

Four studies provided a p-value for comparing SII values in the AA and control groups, all statistically significant [26,27,31,35]. One study performed a three-group comparison (CAA, NCAA, and CG) through a Kruskal-Wallis test, which was statistically significant [32].

Three studies provided a p-value for comparing SII values in the CAA and NCAA, all statistically significant [26,27,31]. One study performed a three-group comparison (CAA, NCAA, and CG) through a Kruskal-Wallis test, which was statistically significant [32].

The random-effect meta-analysis of SII (AA vs. CG) for pediatric populations included five articles (2451 AA and 2355 controls) and resulted in a significant mean difference [95% CI] of 1123.04 [319.52,1926.56] (p<0.01). The I^2^ value was 97.1%. We performed a sensitivity analysis excluding the work by Ortiz-Ley et al. and obtained a significant mean difference [95% CI] of 1460.96 [993.54,1928.37] (p<0.01) (forest plot not shown).

The random-effect meta-analysis of SII (CAA vs. NCAA) included five articles (828 CAA and 1725 NCAA) and resulted in a significant mean difference [95% CI] of 1033.18 [710.92,1355.45] (p<0.001). The I^2^ value was 58.2%.

The forest plot of both meta-analyses is shown in Figure 5.

**Figure 5.**
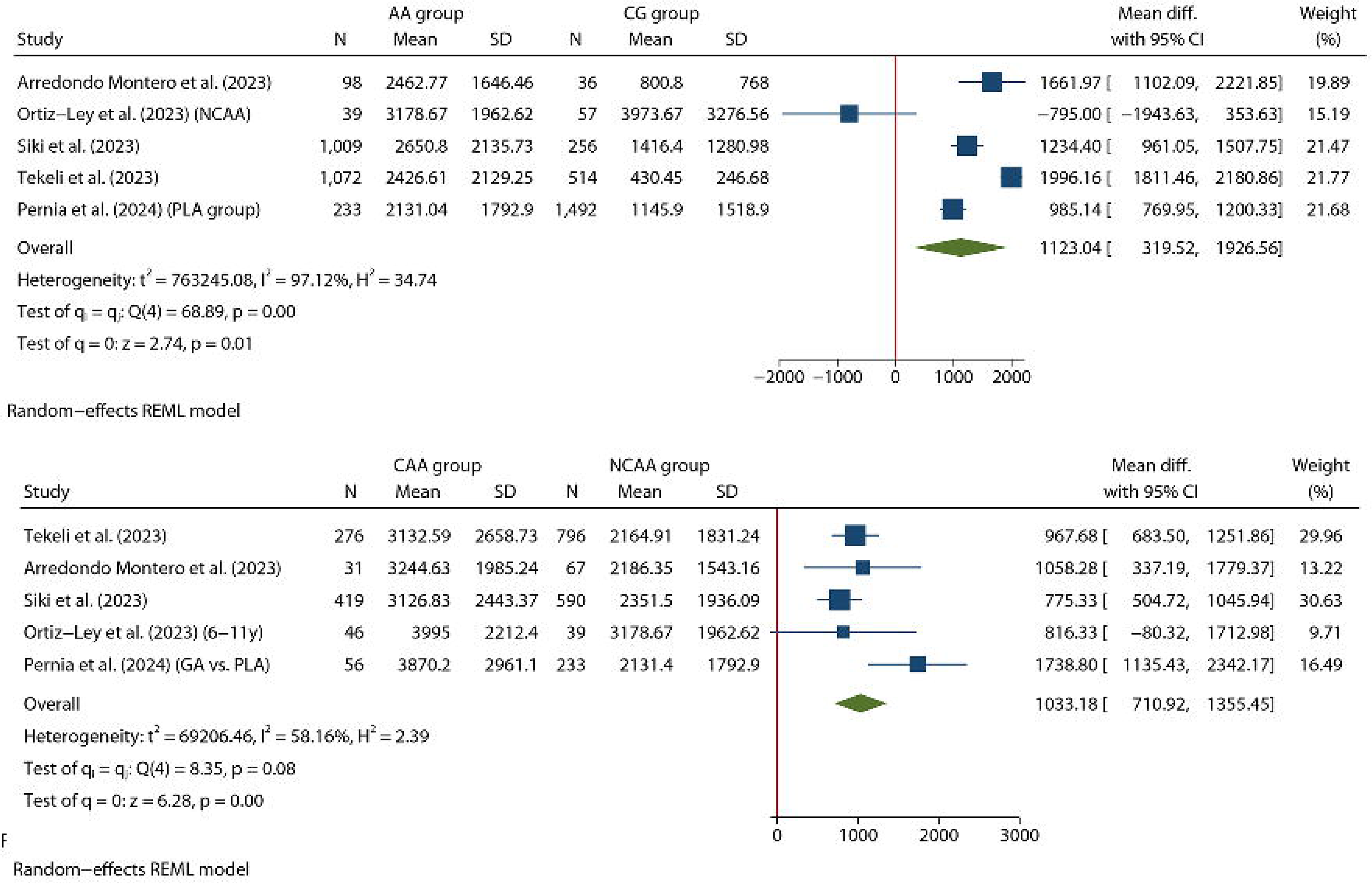
<u>Above</u>: Forest plot of the REML random-effects meta-analysis performed for SII (AA vs. CG) (only pediatric patients included). <u>Bottom</u>: Forest plot of the REML random-effects meta-analysis performed for SII (CAA vs. NCAA) (only pediatric patients included).

## Diagnostic Test Accuracy Meta-Analysis

The DTA meta-analysis (AA vs. CG) yielded a pooled sensitivity and specificity [95% CI] of 81.8 [75.2,86.9] % and 79.9 [68.2,88.1] %. The DTA meta-analysis (NCAA vs. CAA) resulted in a pooled sensitivity and specificity [95% CI] of 72.5 [49.6,87.6] % and 82.5 [65.1,92.2] %. Figure 6 shows the forest plot resulting from both meta-analyses. Figures 7 and 8 show the hSROC curves resulting from both meta-analyses.

**Figure 6.**
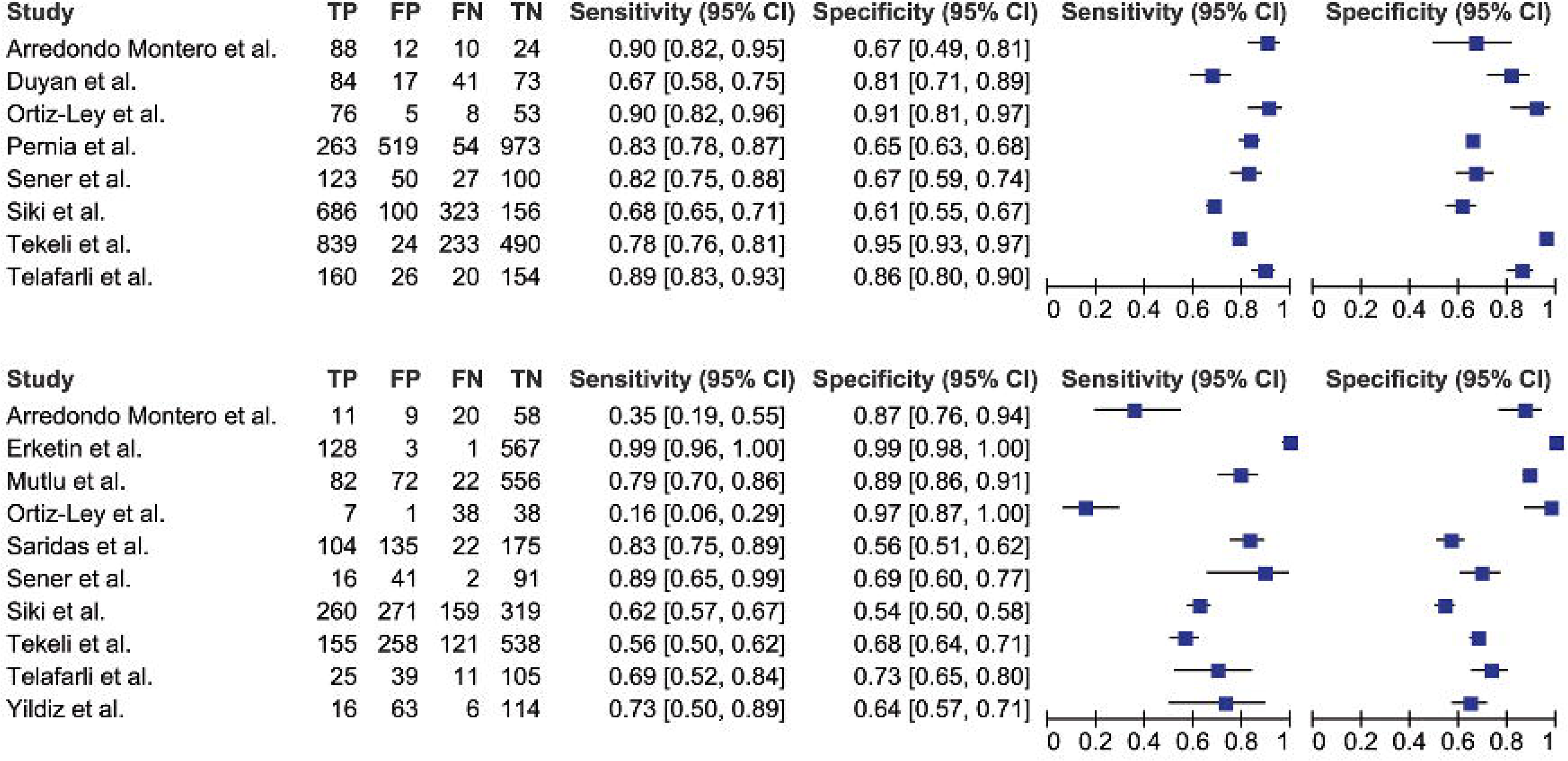
<u>Above</u>: Forest plot of the DTA meta-analysis for SII (AA vs. CG). <u>Bottom</u>: Forest plot of the DTA meta-analysis for SII (CAA vs. NCAA).

**Figure 7.**
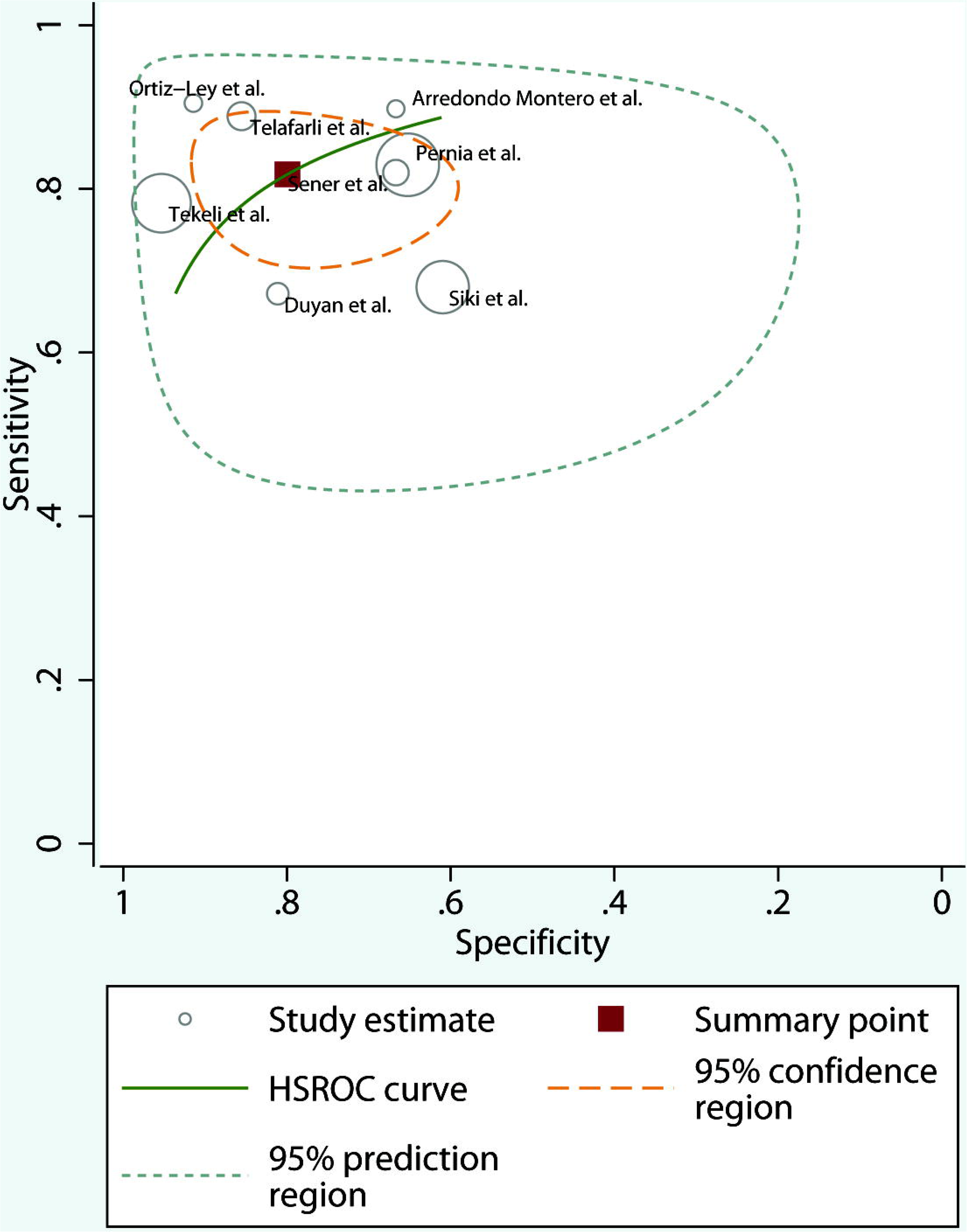
DTA meta-analysis for SII (AA vs. CG). hSROC curve.

**Figure 8.**
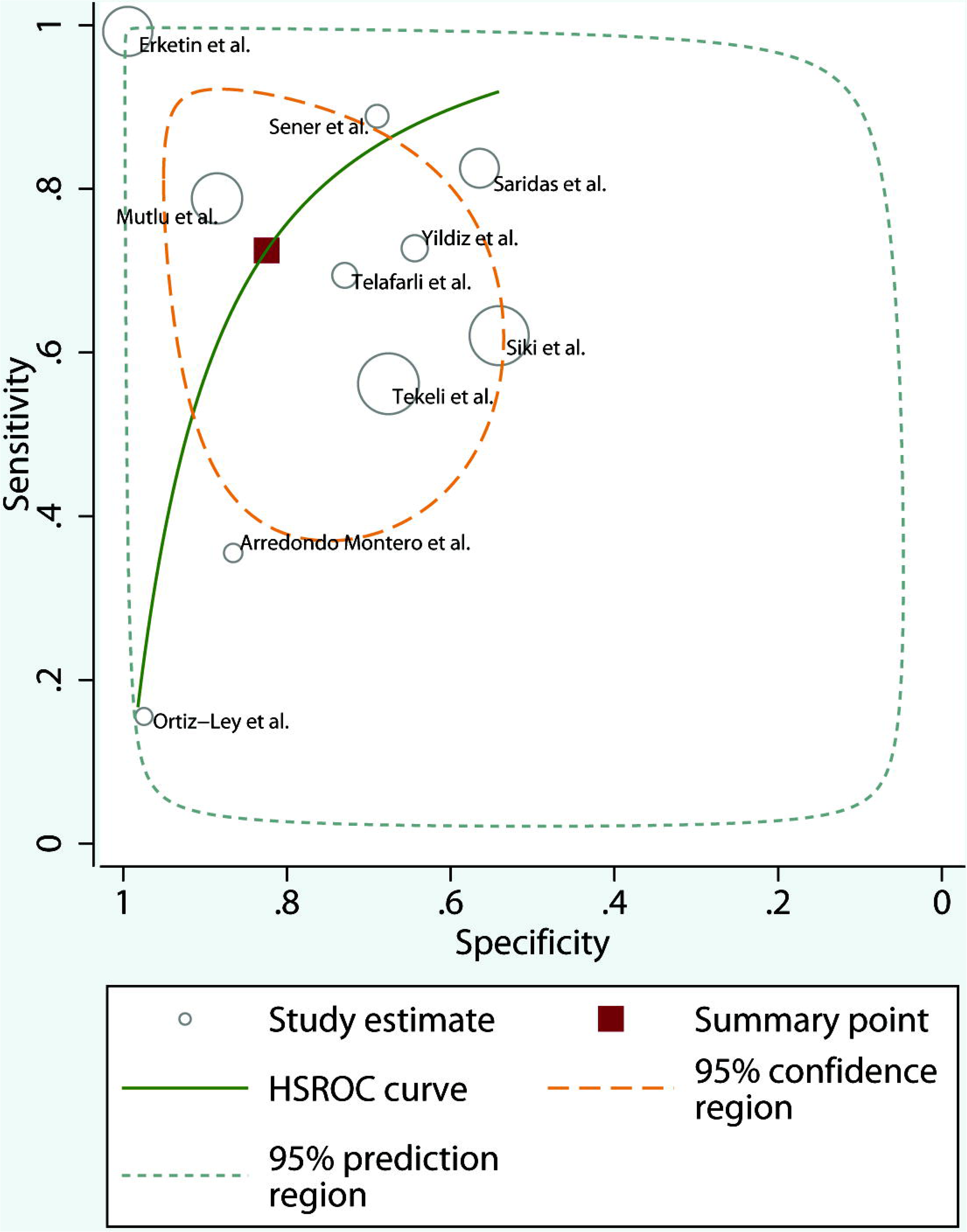
DTA meta-analysis for SII (CAA vs. NCAA). hSROC curve.

## Discussion

The present systematic review and meta-analysis evaluated the role of SII in diagnosing AA and discriminating between NCAA and CAA, consistently demonstrating excellent diagnostic yield in both scenarios. This review also confirmed the usefulness of this index in pediatric populations.

SII is a marker that accurately translates patients’ degree of inflammation and systemic immune response in multiple clinical settings. Concerning AA, the underlying assumption is that AA patients would have a higher degree of inflammation and immune response than patients with NSAP or healthy controls. Similarly, CAA is usually associated with more bacterial translocation, proinflammatory cytokine release, and systemic inflammation than NCAA; therefore, the expected SII levels would be higher. All the papers in this review are consistent and show statistically significant differences in SII values between the AA and CG groups and the CAA and NCAA groups. Similarly, the DTA models showed high pooled sensitivity and pooled specificity values for both AA vs. CG and CAA vs. NCAA analyses.

Concerning the use of hemogram-derived ratios as diagnostic biomarkers, they have multiple advantages that should be considered: 1) CBCs are readily available in virtually any Emergency Department in the world, even in developing countries 2) Their cost in terms of processing and human resources is usually low 3) Results are obtained quickly, which avoids diagnostic delays 4) In many cases patients have a previous baseline CBC, which can be used as an intra-individual reference range. 5) The CBC ranges, in general terms, maintain a reasonably well-defined range of values in humans, with outliers being infrequent. This is a crucial advantage compared to other biomarkers, such as IL-6 or calprotectin, which frequently exhibit outliers.

We consider the creation of these ratios to be a powerful diagnostic tool with great potential. Still, we also believe that changes can be implemented to increase their diagnostic performance: 1) Most authors have conducted their studies assuming that the higher the SII values, the higher the probability of AA diagnosis. However, this need not always be the case. For example, in pediatric populations, the ranges of the different hemogram parameters differ according to the age of the patients [37]. Also, some pathologies, such as pneumonia, may be associated with a higher leukocytosis than AA [38]. It is necessary to consider that some biomarkers may present a quadratic function in diagnostic terms. Giving more value to a specific analytical range and not to the highest extreme values could improve the overall diagnostic performance. 2) For simplification, SII attributes the same value to the three elements that make up the index: TNC, TPC, and TLC. However, the relative weights of the three should be different. The development of statistical and AI models that determine the optimal weighting of each element could also contribute to increasing the diagnostic performance of this index.

Most of the reported diagnostic studies of SII in AA were retrospective. This is a significant limitation, given that the potential presence of selection bias must be assumed in all cases. Also, there is no reliable way to confirm that all patients in the control group did not develop AA (they could have gone to another hospital, for example). Equally, this means that the reported prevalences (of both AA and CAA) may not represent the population where the study was conducted. For example. Şener et al. reported 18 cases of NCAA among 150 AA cases (12% prevalence) [25], but Siki et al. reported 419 cases of CAA among 1009 AA cases (42% prevalence) [31]. The same applies to the sex of the patients included: AA is a pathology with a slight male predominance, and yet some of the studies included (such as that of Ortiz-Ley et al.) [32] have a greater representation of women than men.

Most studies were conducted in the same geographical area (Turkey), making it difficult to extrapolate the results to other populations. Variables such as diet and genetics may influence these results. Therefore, the diagnostic role of SII in AA needs to be validated in different populations.

While there is homogeneity in the definition of cases, there is significant heterogeneity in the definition of the control group. The use of healthy controls or controls without gastrointestinal inflammatory pathology constitutes a critical diagnostic bias since we can erroneously attribute an overdiagnostic capacity to the tool we are evaluating. A healthy person is likely to have a much lower systemic inflammation than someone with an acute gastrointestinal process, even if this is not AA. Studies should be conducted in situations that represent actual clinical practice.

Therefore, the optimal control group should comprise patients with a formal suspicion of AA in whom this diagnosis is reliably excluded (NSAP patients). On the other hand, excluding septic patients, considered by some authors, may also bias the results if the sepsis is in the context of an AA since these patients present the pathology and, in our opinion, should be considered in the diagnosis.

Lastly, the control group from Guo et al. [35], defined as chronic appendicitis, is not representative of the object of study of this review and was, therefore, excluded from all meta-analytic models performed.

Ortiz-Ley et al. reported using a control group of healthy patients and considered a CBC from the fifteen days before inclusion valid. This does not allow for adequate control of confounding factors, as patients may have had undocumented inflammatory processes during that period. Interestingly, this study is the only one reporting high values for the healthy outpatient group. This is also in line with the leave-one-out model for the first random effects meta-analysis we performed (AA vs. CG), where the exclusion of this study significantly improved the model. Likewise, when we excluded such work from the meta-analytic model of pediatric populations (AA vs. CG), the model improved ostensibly.

We also found an important source of heterogeneity in the studies included in this review regarding the inclusion and exclusion criteria. Some studies do not explicitly state their exclusion criteria, which is a significant limitation. CBC is a sensitive test, easily altered by any inflammatory or infectious process. Therefore, ensuring that patients strictly comply with the detailed inclusion and exclusion criteria is essential. Future studies should imperatively homogenize these criteria.

Finally, the analyses performed show that it is also a potentially useful diagnostic tool in the pediatric subgroup. The present study presents essential strengths, such as the robust methodology based on the PRISMA-DTA guidelines and the meta-analytical models used. On the other hand, it has significant limitations: 1) The limited number of articles and patients included, 2) The retrospective nature of most included studies, 3) the high heterogeneity found in the meta-analytical models performed, and 4) The limitations inherent to the inferential statistical procedures used.

In conclusion, SII is a biomarker with excellent diagnostic performance for AA and for discriminating NCAA from CAA in adult and pediatric populations. The retrospective nature of most reported studies and their limited geographic variability justify new prospective multicenter studies to validate these findings.

## Supporting information

Supplementary File 1. PRISMA-DTA checklist.

Supplementary File 2. Inclusion and exclusion criteria.

## Data Availability

The data used to carry out this systematic review are available upon request from the authors.

