## Supplementary File 2. Inclusion and exclusion criteria. for "Diagnostic Performance of Systemic-Immune Inflammation Index for Overall and Complicated Acute Appendicitis: A Systematic Review and a Diagnostic Test Accuracy Meta-Analysis"

**Supplementary File 1. Inclusion and Exclusion Criteria**

**Inclusion criteria**

-Pilot prospective or retrospective observational original clinical studies measuring the diagnostic performance of systemic-immune inflammation index compared to the reference standards for diagnosing appendicitis and for discriminating between complicated and uncomplicated appendicitis.

-Original diagnostic validation studies measuring the diagnostic performance of systemic-immune inflammation index compared to the reference standards for diagnosing appendicitis and the discrimination between complicated and uncomplicated appendicitis.

**Exclusion criteria**

-Case reports

-Duplicate or overlapping studies.

-Reviews, systematic reviews, consensus guidelines.

-Languages other than English or Spanish.

-Studies with no surgical intervention.

-Studies with no population of interest.

-Studies conducted in immunocompromised patients.

-Studies conducted in patients with metastatic neoplastic disease and invasive abdominal neoplastic disease.

-Studies conducted on patients with hematological diseases.

-Studies conducted in pregnant patients.
