## Supplementary material for "Diagnostic Performance of Systemic-Immune Inflammation Index for Overall and Complicated Acute Appendicitis: A Systematic Review and a Diagnostic Test Accuracy Meta-Analysis": Table 1.

| **Author** | **Country** | **Study design** | **Age (Range)** | **Sex M/F** | **Total*N*** | ***N* in AA** | ***N* in CG** | **SII AA** | **SII CG** | **P**  **(CG vs AA)** | **P (NCAA vs CAA)** | **Cut-off U/mL** | **AUC [95% CI]** | **Sensitivity (%)** | **Specificity (%)** |
| --- | --- | --- | --- | --- | --- | --- | --- | --- | --- | --- | --- | --- | --- | --- | --- |
| Duyan et al. (2022) | Turkey | Retrospective | 40.2 (15.1)y^1^ | 181/166 | 347 | 125 | 222  NSAP: 90  HC: 132 | AA: 1936.43 (1276.06)^1^ | NSAP: 950.69 (853.09)^1^  NIAP: 713.89 (457.58)^1^ | AA vs. NSAP: <0.001  AA vs. NIAP: <0.001 | - | AA vs. NSAP: 1218.97 | AA vs. NSAP: 0.80 [0.74-0.85] | AA vs. NSAP: 67.2 | AA vs. NSAP: 81.1 |
| Şener et al. (2022) | Turkey | Retrospective | AA: 33.47 (11.01)y^1^  CG: 35.67 (12.23)y^1^ | 166/134 | 300 | 150  NCAA: 132  CAA: 18 | NSAP: 150 | AA: 1759.62 (1263.92)^1^ | NSAP: 979.96 (1032.33)^1^ | AA vs. NSAP: <0.001 | - | AA vs. NSAP: 840.13  NCAA vs CAA: 1782.94 | NSAP vs AA: 0.764 [0.709-0.819]  NCAA vs CAA: 0.826 [0.744-0.909] | AA vs. NSAP: 82  NCAA vs. CAA: 88.9 | AA vs. NSAP: 66.7  NCAA vs. CAA: 68.9 |
| Tekeli et al. (2023)^a^ | Turkey | Retrospective | AA: 11.52 (3.74)y^1^  CG: 10.35 (2.98)y^1^ | 1023/563 | 1586 | 1072  NCAA: 796  CAA: 276 | HC: 514^b^ | AA: 2426.61 (2129.25)^1^  NCAA: 2164.91 (1831.24)^1^  CAA: 3132.59 (2658.73)^1^ | HC: 430.45 (246.68)^1^ | AA vs. HC: <0.001 | NCAA vs. CAA: <0.001 | AA vs. HC: 923  NCAA vs. CAA: 2358.03 | AA vs. HC: 0.927  NCAA vs. CAA: 0.646 | AA vs. HC: 78.3  NCAA vs. CAA: 56.4 | AA vs. HC: 95.33  NCAA vs. CAA: 67.67 |
| Arredondo Montero et al. (2023)^a^ | Spain | Prospective | AA: 9.69 (3.08)y^1^  CG: 10.65 (2.55)y^1^ | 81/53 | 134 | 98  NCAA: 67^c^  CAA:31^c^ | NSAP: 36 | AA: 2381.85 (1409.14-3597.33)^2^  AA: 2462.77 (1646.46)^4^  NCAA: 2199.6 (1161.33-3198.11)^2,c^  NCAA:2186.35 (1543.16)^4^  CAA: 3195.5 (1992-4546.39)^2,c^  CAA: 3244.63 (1985.24)^4^ | NSAP: 696.34 (355.67-1350.38)^2^  NSAP: 800.8 (768)^4^ | AA vs. NSAP: <0.0001 | NCAA vs. CAA: 0.01^c^ | AA vs. NSAP: 890  NCAA vs. CAA: 4217^c^ | AA vs. NSAP: 0.85 [0.78-0.92]  NCAA vs. CAA: 0.66 [0.54-0.77]^c^ | AA vs. NSAP: 89.8  NCAA vs. CAA: 35.5^c^ | AA vs. NSAP: 66  NCAA vs. CAA: 86.6^c^ |
| Mutlu et al. (2023) | Turkey | Retrospective | NCAA: 33.2 (14.1)y^1^  CAA: 34.3 (13.4)y^1^  CG: 34.8 (13.6)y^1^ | 810/636^d^ | 1446^d^ | 732  NCAA: 628  CAA. 104 | NIAP: 714 | NCAA: 1797.8 (430.2)^1^  CAA: 2514.3 (890.9)^1^ | NIAP: 757.1 (303.6)^1^ | AA vs. NIAP: <0.001 | NCAA vs. CAA: <0.001 | NCAA vs. CAA: 1989.2 | NCAA vs. CAA: 0.809 [0.768-0.844] | NCAA vs. CAA: 78.44 | NCAA vs. CAA: 88.52 |
| Telafarlı et al. (2023) | Turkey | Retrospective | AA: 31.12 (9.2)y^1^  CG: 29.48 (7.35)y^1^ | 251/109 | 360 | 180  NCAA: 144  CAA: 36 | NSAP: 180 | AA: 2438.88 (1987.14)^1^  NCAA: 1773.8 (1112.83)^1^  CAA: 5099.19 (2456.82)^1^ | NSAP: 1282.9 (1193.33)^1^ | AA vs. NSAP: <0.001 | NCAA vs. CAA: <0.001 | AA vs. NSAP: 2726.26  NCAA vs. CAA: 1239.77^e^ | AA vs. NSAP: 0.75 [0.7-0.8]^e^  NCAA vs. CAA: 0.927 [0.889-0.965]^e^ | AA vs. NSAP: 88.9^e^  NCAA vs. CAA: 70.6^e^ | AA vs. NSAP: 85.4^e^  NCAA vs. CAA:72.8^e^ |
| Yıldız et al. (2023) | Turkey | Retrospective | AA: 38.5 (14.7)y^1^  CG: 41.3 (15.5)y^1^ | 125/95 | 220 | 199  NCAA: 177  CAA: 22 | NA: 21 | AA: 1735.1 (823.8-1989.25)^2^  AA: 1516.05 (870.30)^4^  NCAA: 1207 (571.5-2089)^2^  NCAA: 1289.17 (1134.24)^4^  CAA: 2514.5 (1132.25-5388)^2^  CAA: 3011.42 (3372.63)^4^ | NA: 812.1 (662.1-1012.32)^2^  NA: 828.84 (278.44)^4^ | AA vs. NA: 0.002 | NCAA vs. CAA: <0.001 | NCAA vs. CAA: 1465 | NCAA vs. CAA: 0.786 [0.586-0.820] | NCAA vs. CAA: 72.7 | NCAA vs. CAA: 64.2 |
| Siki et al. (2023)^a^ | Turkey | Retrospective | CG: 11 (0-17)y^2,f^  AA: 10 (0-17)y^2,f^ | 784/481 | 1265 | 1009  NCAA: 590  CAA: 419 | NA: 256 | AA: 2489.5 (1293-4169.9)^2^  AA: 2650.8 (2135.73)^4^  NCAA: 2111.8 (1168.7-3774)^2^  NCAA:2351.5 (1936.09)^4^  CAA: 2873.9 (1611-4895.6)^2^  CAA: 3126.83 (2443.37)^4^ | NA: 1177 (677-2395.2)^2^  NA: 1416.4 (1280.98)^4^ | AA vs. NA: <0.001 | NCAA vs. CAA: <0.001 | AA vs. NA: 1630.1  NCAA vs. CAA: 2296 | AA vs. NA: 0.69 [0.664-0.716]  NCAA vs. CAA: 0.595 [0.564-0.625] | AA vs. NA: 68  NCAA vs. CAA: 62 | AA vs. NA: 61  NCAA vs. CAA: 54 |
| Ortiz-Ley et al. (2023)^a^ | Mexico | Retrospective | 9 (5-12.5)y^2^ | 156/221 | 377 | 188^g^  NCAA: 94  (3-5 years, n=21; 6-11 years, n=39; >12 years, n=34)  CAA: 94  (3-5 years, n=21; 6-11 years, n=46; >12 years, n=27) | CG: 189^g, h^  (3-5 years, n=63; 6-11 years, n=57; >12 years, n=69) | 3-5 years:  NCAA: 1685 (1128-2250)^2,h^  CAA: 3560 (2300-4830)^2,h^  6-11 years:  NCAA: 3266 (1860-4410)^2,h^  NCAA:3178.67 (1962.62)^4^  CAA: 3980 (2558-5447)^2,h^  CAA: 3995 (2212.4)^4^  >12years:  NCAA: 2510 (1220-4490)^2,h^  CAA: 3527 (2327-5780)^2,h^ | 3-5 years:  CG: 1940 (1130-2660)^2,h,i^  6-11 years:  CG: 3631 (1990-6300)^2,h,i^  CG: 3973.67 (3276.56)^4^  >12years:  CG: 4100 (2860-7020)^2,h,i^ | - | 3-5 years:  <0.001  6-11 years:  <0.001  >12years:  <0.001  (three-group comparison values: NCAA, CAA, and CG) | 3-5 years:  AA vs. CG: 4970  NCAA vs. CAA: 2250  6-11 years:  AA vs. CG: 1240  NCAA vs. CAA: 7560  >12years:  AA vs. CG: 1000  NCAA vs. CAA: 5610 | 3-5 years:  AA vs. CG: 0.97 [0.93-0.99]  NCAA vs. CAA: 0.8 [0.6-0.9]  6-11 years:  AA vs. CG: 0.97 [0.93-0.98]  NCAA vs. CAA: 0.61 [0.47-0.72]  >12years:  AA vs. CG: 0.92 [0.85-0.96]  NCAA vs. CAA: 0.85 [0.77-0.91] | 3-5 years:  AA vs. CG: 92  NCAA vs. CAA: 85  6-11 years:  AA vs. CG: 91  NCAA vs. CAA: 15  >12years:  AA vs. CG: 86  NCAA vs. CAA: 29 | 3-5 years:  AA vs. CG: 92  NCAA vs. CAA: 76  6-11 years:  AA vs. CG: 91  NCAA vs. CAA: 97  >12years:  AA vs. CG: 86  NCAA vs. CAA: 94 |
| Ertekin et al. (2023) | Turkey | Retrospective | NCAA: 29 (11-75)y^3^  CAA: 30 (18-70)y^3^ | 455/244 | 699 | 699  NCAA: 570  CAA: 129 | - | NCAA: 863 (60-8576)^3,j^  NCAA: 2590.5 (1390.19)^5^  CAA: 16022 (4641-773696)^3,j^  CAA: 202595.25 (148617.15)^5^ | - | - | NCAA vs. CAA: <0.001 | NCAA vs. CAA: 5703.3 | NCAA vs. CAA: 0.999 [0.999-1] | NCAA vs. CAA: 99.2 | NCAA vs. CAA: 99.5 |
| Pernia et al. (2024)^a^ | Spain | Retrospective | 10.1 (7.7-11.9)y^2^ | 1101/708 | 1809 | 317  PLA: 233  GA: 56  PA: 22  Peritonitis/abscess: 6 (authors clarified that these six patients don’t belong to the GA/PA group)^k^ | CG: 1492  NA: 19 | PLA: 2131.4 (1792.9)^1,c^  GA: 3870.2 (2961.1)^1,c^  PA: 3487.8 (1745.8)^1,c^  Peritonitis: 2904.2 (1162.0)^1,c^ | CG: (NA + NSAP): 1145.9 (1518.9)^1,c^ | - | - | AA vs. NSAP: 923 | AA vs. CG: 0.788 [0.767-0.807]  AA vs. CG: 0.741 [0.720 - 0.763] | AA vs. CG: 83.1^c^ | AA vs. CG: 65.2^c^ |
| Saridas et al. (2024) | Turkey | Retrospective | NCAA: 39.21 (31.09)y^1^  CAA: 35.14 (13.81)y^1^ | 239/197 | 436 | 436  NCAA: 310  CAA: 126 | - | NCAA: 1218.21 (634.21)^1^  CAA: 1886.54 (850.62)^1^ | - | - | NCAA vs. CAA: <0.001 | NCAA vs. CAA: 1647.01 | NCAA vs. CAA: 0.742 [0.7-0.79] | NCAA vs. CAA: 82.6 | NCAA vs. CAA: 56.3 |
| Guo et al. (2024)^a^ | China | Retrospective | AA: 9.75 (2.14)y^1^ CRA: 9.48 (2.05)y^1^ | 55/49 | 104 | 51 | CRA: 53 | AA: 660.41 (80.34)^1^ | CRA: 605.27 (78.25)^1^ | AA vs. CRA: <0.001 | - | AA vs. CRA: AUC combines SII and PAS; the authors do not provide the AUC for SII alone. | - | - | - |

**Table 1. Summary of publications included in this review.**

**AA:** Acute appendicitis group, **CG**: Control group, **NCAA**: Non-complicated acute appendicitis, **CAA:** Complicated acute appendicitis, **NIAP**: Non-inflammatory abdominal pain; **NS**: non-statistically significant **PLA**: Phlegmonous appendicitis; **GA**: Gangrenous appendicitis; **PA**: perforated appendicitis; **NSAP:** Non-surgical abdominal pain (Suspected AA subsequently excluded in the Emergency Department); **HC**: Healthy controls; **NA**: Negative Appendectomies; **SII**: Systemic-immune inflammation index; **SIRI**: systemic-immune response index; **y**: years; **CRA**: Chronic Appendicitis; **PAS**: Pediatric Appendicitis Score.

**1**: Mean (standard deviation); **2:** Median (Interquartile range); **3:** Median (range); **4:** Mean (standard deviation) calculated from Median (Interquartile range); **5:** Mean (standard deviation) calculated from Median (Range)

***a****: Study conducted on Pediatric patients;* ***b****:* *In some parts of Tekeli et al. manuscript, 541 patients are reported (e.g., in the abstract), and in others, 514 (e.g., in Table 1).* *We used the value 514, obtained when the number of males and females in Table 1 for this group was summed. The corresponding author confirmed that 514 was the correct number;* ***c****: Data not available in the original article, provided by the authors;* ***d****: The manuscript by Mutlu et al. has inconsistencies in the number of patients: authors report 628 patients with NCAA; 104 patients with CAA and 714 controls (total n = 1446) but the total sample size reported in the main text and the abstract is 1456 patients. On the other hand, the number of males in the control group should correspond to 55.6% (according to the authors), but only 109 patients out of 714 males are reported. The same occurs in the CAA group (Authors report 58.5% of males, but only 24 out of 104 males figure in Table 1). We have chosen to recalculate the number of male patients based on the percentage provided and to include 1446 rather than 1456 patients in the table;* ***e****: The authors (Telafarlı et al.) propose a higher cut-off for diagnosing AA (AA vs. NSAP) (2726.26) than for diagnosing CAA (NCAA vs. CAA) (1239.77). This is statistically inconsistent, as the SII values are higher in both groups for the NCAA vs. CAA comparison than for the AA vs. NSAP comparison. One explanation could be that the NSAP vs. CAA comparison was made instead of NCAA vs. CAA, although the authors do not make this explicit. Also, in the case of SII (NSAP vs. AA), a sensitivity of 88.9 and a specificity of 85.4 are reported, much higher than expected for an AUC of 0.75, while in the case of SII (NCAA vs. CAA), the opposite happens: a sensitivity of 70.6 and a specificity of 72.8 are reported for an AUC of 0.927 when much higher values would be expected;* ***f****: In the Study by Siki et al. age data on table 1 is reported as median (interquartile range). Nevertheless, the reported data is more consistent with the median (range);* ***g****: In the study conducted by Ortiz-Ley et al., there are inconsistencies in the number of patients. The authors reported 189 patients in the non-AA group, but the sum of the three age groups for this category (non-AA) is 190 (n=63 + n=58 +n=69). Same occurs for the NCAA (n=21 + n=39 + n=34) and CAA (n=21 + n=45 +n=27) groups (sum = 187, expected =188). We contacted the authors, and the numbers included in the table are correct, as reported by the corresponding author after contacting her;* ***h****: Ortiz-Ley et al reported SII as (x10^6^). SII is a ratio and, therefore, dimensionless, although in the literature, it is always reported after calculation with hemogram parameters quantified as 1x10^9^/L. The relevant conversion to these values has been performed (multiplying by 1000), and the results are consistent with the above literature and have been confirmed with the authors;* ***i****: The CG for Ortiz-Ley et al. is defined as “**patients without AA who were seen in pediatric outpatients without a diagnosis of appendicitis.”. After contacting the authors, they clarified that these patients were included during pre-surgical consultations for scheduling outpatient surgery and did not have any inflammatory or infectious process at the time of inclusion in the study.;* ***j****:* *Although Ertekin et al. values are supposed to be reported as median (IQR), values are more compatible with median (range);* ***k****: Pernia et al. reported a discrepancy between sample size per group in the main text and Figure 1. We included values from Figure 1 since the true positives, true negatives, false positives, and false negatives matched these values.*
